# Linking objective measures of physical activity and capability with brain structure in healthy community dwelling older adults

**DOI:** 10.1101/2021.01.28.21250529

**Authors:** Anne-Marthe Sanders, Geneviève Richard, Knut Kolskår, Kristine M. Ulrichsen, Tobias Kaufmann, Dag Alnæs, Dani Beck, Erlend S. Dørum, Ann-Marie G. de Lange, Jan Egil Nordvik, Lars T. Westlye

## Abstract

Maintaining high levels of daily activity and physical capability have been proposed as important constituents to promote healthy brain and cognitive aging. Studies investigating the associations between brain health and physical activity in late life have, however, mainly been based on self-reported data or measures designed for clinical populations. In the current study, we examined cross-sectional associations between physical activity, recorded by an ankle-positioned accelerometer for seven days, physical capability (grip strength, postural control, and walking speed), and neuroimaging based surrogate markers of brain health in 122 healthy older adults aged 65-88 years. We used a multimodal brain imaging approach offering two complementary structural MRI based indicators of brain health: white matter diffusivity and coherence based on diffusion tensor imaging and subcortical and global brain age based on brain morphology inferred from T1-weighted MRI data. The analyses revealed a significant association between global white matter fractional anisotropy (FA) and walking speed, indicating higher white matter coherence in people with higher pace. We also found a significant interaction between sex and brain age on number of daily steps, indicating younger-appearing brains in more physically active women, with no significant associations among men. These results provide insight into the intricate associations between different measures of brain and physical health in old age, and corroborate established public health advice promoting physical activity.

## 1. Introduction

Magnetic resonance imaging (MRI) studies have revealed substantial age-related changes in the human brain, including cortical and subcortical atrophy, ventricular enlargement, and white matter alterations (Fjell et al., 2009; Garde et al., 2000; Westlye et al., 2010; Ylikoski et al., 1995). While group analysis suggests robust effects, studies have revealed considerable heterogeneity across individuals and brain structures (Allen et al., 2005; Fjell et al., 2013; Raz et al., 2010; Sexton et al., 2014; Tucker & Stern, 2011). As further understanding of the complexity and biological basis of brain and cognitive aging evolves, more targeted preventive measures and innovative models of geriatric care is expected to be developed as part of public health programs.

Physical health indicators, such as daily activity level, balance, walking speed, and hand-grip strength, are associated with healthy aging (Kuh, 2007; Vermeulen et al., 2011), and represent a putative malleable moderator of brain aging. A number of studies have reported positive associations between these indicators and the volume and integrity of brain white and gray matter structures (Bherer et al., 2013; Erickson et al., 2014), including the subcortical areas hippocampus (Erickson et al., 2014; Hamer et al., 2018) and the cerebellum (Chen et al., 2015; Surgent et al., 2019). However, most previous studies are limited by the use of self-report measures of physical activity, including mainly clinical populations or assessment tools primarily designed for clinical populations (Demnitz et al., 2018), which may not be sensitive to detect relevant individual differences among healthy adults. Among the exceptions, a study of healthy adults aged >80 years reported an association between high levels of accelerometer-measured daily steps and global fractional anisotropy (FA) (Tian et al., 2015), which is an indicator of overall white matter coherence based on diffusion tensor imaging (DTI). An association has also been found between temporal and parahippocampal white matter FA and accelerometer-measured daily steps in low-active adults aged 60-78 years reporting less than 150 minutes of moderate physical activity per week (Burzynska et al., 2014). Sex differences in beneficial effects of physical activity on brain health have also been reported. For example, higher number of accelerometer-measured daily walking steps was related to larger surface area in subregions of the hippocampus in women but not in men (Varma et al., 2016).

Recent advances in MRI analysis and machine learning have shown that complex, multidimensional brain imaging data can be aggregated into a sensitive, unitary estimate of brain aging (Cole & Franke, 2017; Franke et al., 2010). Estimated high brain age compared to chronological age (brain age gap; BAG) in older adults has been suggested to indicate incipient neurodegeneration (Franke & Gaser, 2019) and increased risk of dementia (Wang et al., 2019). Brain age prediction and similar data-driven approaches provide innovative imaging based surrogate markers of brain health, with promising potential to translate complex neuroscience data to more comprehendible public health guidelines, thus bridging the gap between public health and science. The World Health Organization describes improved understanding of analytical approaches for healthy aging a priority of action, including development of biomarkers related to healthy aging (World Health Organization, 2015). To our knowledge, only a few prior studies have investigated the link between BAG and physical activity and capabilities (Cole et al., 2018; Steffener et al., 2016), and only one included objective measures like hand-grip strength and walking speed (Cole & Franke, 2017). Overall, the evidence linking objective measures of physical health and indicators of brain health is conflicting or lacking, both with regard to white matter microstructure (Kilgour et al., 2014; Wassenaar et al., 2019) and BAG based on gray matter morphology. In addition, it is not clear to which degree T1-weighted structural MRI measures and DTI-based markers of white matter coherence show differential sensitivity to physical health in ageing men and women.

The purpose of the current study was to examine the association between objective measures of physical activity level and capability, and two complementary structural MRI based indicators of brain health: white matter coherence based on DTI and BAG based on brain morphology. We included 122 (62% women) community-dwelling healthy middle-aged and older adults aged 65-88 years. Physical activity was measured using ankle-worn accelerometer across on average 7 days (range 3 to 9) and physical capabilities were operationalized through grip strength, walking speed, and postural control as a measure of balance. Based on recent implementations and the use of an independent training set, we estimated individual brain age using global and subcortical gray matter from T1-weighted MRI (de Lange & Cole, 2020; de Lange et al., 2019; Kaufmann et al., 2019). We used an independent training set comprising MRI data from 2407 healthy individuals for brain age prediction, and applied the cross-validated models in our unseen test set. In addition, to complement the brain age approach we included DTI-based global fractional anisotropy (FA) and mean diffusivity (MD).

Based on the putative close link between brain and physical health, we hypothesized that level of physical activity, hand-grip strength, walking speed, and postural control, would be associated with BAG, both estimated using global and subcortical measures, and global FA and MD. Moreover, based on previous evidence we tested for sex differences in the associations between physical health indicators and brain MRI measures, as well as differences in the sensitivity to the physical health indicators between the brain gray and white matter measures.

## 2. Materials and methods

### 2.1. Participants

Data collection was performed as an integrated part of the prospective StrokeMRI study aiming at identifying predictors of brain and cognitive health, aging and stroke rehabilitation (Dørum et al., 2016; Dørum et al., 2017; Richard et al., 2018). Briefly, healthy adults were recruited from the Oslo area in Norway through advertisements in local papers and by word of mouth, and screened for eligibility through a standardized phone interview before inclusion. Participants reporting counter-indication for MRI, history of severe psychiatric or neurological conditions, for example epilepsy, brain tumour or head trauma with loss of consciousness for more than two minutes, and/or alcohol/drug abuse, were excluded.

All participants (n=341, 18-94 years) completed demographic information, standardized questionnaires, including for example mood, personality, and sleep habits, a comprehensive cognitive test battery (Richard et al., 2018), clinical and medical assessments, and multimodal structural and functional MRI. In the current study we included participants aged 65 years or older who additionally completed standardized physical tests and accelerometer assessment.

Among a total of 131 eligible participants, nine were excluded due to poor quality on T1-weighted data (n=5), not completing the MRI protocol (n=3), or not completing the tests of physical capability and physical activity (n=1), reducing the number of participants to n=122, with an average age of 71.4 years (SD=4.61, 64-88 years, 62% women). No participants scored below 24 on the Mini-Mental State Examination (MMS-E) (Folstein et al., 1975). For the current sample, the median interval between MRI and the neuropsychological and physical tests was 11 days (IQR= 14.75 days).

The study was completed in accordance with the Helsinki Declaration and approved by the Regional Committees for Medical and Health Research Ethics for the South-Eastern Norway (REK approvals 2014/694, 2015/1282). All participants provided written informed consent. The participants received compensation for participation in the study.

### 2.2. Cohort used for training set in brain age prediction

The training sample consisted of data from 2407 healthy individuals aged 18-94 years from six different samples (Supplementary Table 1, Supplementary Fig. 1). The participants had been recruited and screened in line with local procedures (Supplementary Table 2), in general ensuring that the participants did not have any contraindications for MRI, or a previous or current serious neurological or psychiatric condition, drug abuse, or head trauma.

### 2.3. Measures of physical activity and capability

Four measures of physical activity and capability were considered, including daily step count, postural control, walking speed, and hand-grip strength.

A calibrated StepWatch Activity Monitor (The Modus StepWatchTM3 Activity Monitor) assessed physical activity level. The StepWatch was placed above the ankle and calibrated according to the participant’s height and weight. The instructions for use included wearing the monitor for seven consecutive days, all waking hours, while conducting regular daily activities. Exceptions included bathing or showering. The StepWatch records number of steps taken per one-minute sampling period from one leg. Results were doubled to capture the number of steps taken by each participant (Doherty et al., 2017). According to regular procedures for calculation of activity level, a day was considered valid if consisting of minimum 600 minutes recordings (Mâsse et al., 2005). Non-wear time was defined as >90 consecutive minutes of zero counts. A deviation from this rule was if there was a period of maximum two minutes with more than zero counts within a 90 minutes period with non-wear. The 30 minutes before and after this interruption needed to be consistently zero counts. A minimum of three valid days of data per participant were required for the analysis (Jefferis et al., 2014; Mudge et al., 2010). Valid StepWatch data was available from all 122 participants, who in average wore the StepWatch for 859 minutes per day (Standard Deviation [SD] = 66.4), ranging between 3 and 9 days, with a median of 7 days.

Postural control, registered through measurement of centre of pressure, was assessed using a force plate (BTrackS Balance Tracking System Inc. San Diego, CA, USA, 25 Hz). The protocol for measuring centre of pressure was based on recommendations from Low and colleagues (2017), with double leg stance and 60 second durations of each trial. The balance board was turned towards and 20 cm away from a wall to prevent the participant from turning his or her head or eyes during testing. The participants were asked not to voluntarily move, to keep their hands on their hips, and to position their feet with shoulder-width separation. The data were filtered with a dual-order low pass Butterworth filter with a cut-off frequency of 4 Hz. The average of three trials with eyes closed were used to calculate the mean velocity in cm per second.

Comfortable gait speed was assessed using Timed 10-Meter Walk Test (10 MWT) (Bohannon et al., 1996; Studenski et al., 2011; Wolf et al., 1999). The middle six metres were timed, excluding the effect of acceleration and deceleration, and converted to meter per second. Dominant hand grip strength, expressed in kilograms (kg), was measured with a Jamar hand dynamometer (Sammons Preston Inc., Bolingbrook, IL) with the use of the second handle position for optimal performance. Standard procedure for hand-grip strength test was followed, with the participants in a seated position and elbow-joint fixed in a 90 degrees flection (Mathiowetz et al., 1984). For both walking speed and hand-grip strength, the average of three trials were considered.

### 2.4. MRI acquisition, processing and analysis

Participants were scanned with a 3T General Electric (GE) 750 Discovery MRI scanner using identical sequences and a 32-channel head coil at Oslo University Hospital, Norway. Cushioning was used to minimize head motion.

T1-weighted images were obtained using an inversion recovery-fast spoiled gradient echo (BRAVO) sequence with echo time (TE) = 3.18 ms, repetition time (TR) = 8.16 ms, field of view (FOV) = 256 mm x 256 mm, flip angle (FA) = 12°, and voxel size (VS) = 1×1×1 mm. The images were acquired in sagittal plane, and it took 4.43 minutes to acquire 188 slices. A detailed description of image acquisition for the training samples is provided in Supplementary material Table 3.

DTI data were obtained using an echo planar imaging (EPI) sequence with 60 unique directions, b-value of 1000 s/mm^2^, TE= 83.1 ms, TR = 8150 ms, FA= 90°, FOV = 128 x 128 mm, 2 mm isotropic voxels, and 5 b=0 volumes. Scan time was 8:58 minutes. 7 b=0 volumes with reversed phase-encoding direction were also obtained.

FreeSurfer 5.3 (http://surfer.nmr.mgh.harvard.edu/) was used for automated surface-based morphometry and subcortical segmentation of the T1-weighted images, providing measures of cortical thickness, area, and volume, as well as the volumes of subcortical structures. Technical details of the procedures are described in prior publications (Dale et al., 1999; Fischl et al., 2002). A visual quality-check of reconstructions and subsequent correction, if required, was performed for the StrokeMRI data. The training samples, consisting of data from multiple sites, were all quality checked using FreeSurfer’s Euler number as a proxy (Rosen et al., 2018).

DTI data were processed using Oxford Centre for Functional Magnetic Resonance Imaging of the Brain (FMRIB) Software Library (FSL) (https://fsl.fmrib.ox.ac.uk/fsl). Using topup (https://fsl.fmrib.ox.ac.uk/fsl/fslwiki/topup) and eddy (https://fsl.fmrib.ox.ac.uk/fsl/fslwiki/eddy), we corrected for motion, eddy currents, and geometrical distortions (based on the b=0 volumes collected with a reversed phase-encoding direction) in an integrated stream (Andersson & Sotiropoulos, 2016) which also includes identification and replacement of outlier slices (Andersson et al., 2016). The resulting corrected datasets were used to estimate FA and MD using *dtifit* in FSL.

Further processing was performed using Tract-Based Spatial Statistics (TBSS) (Smith et al., 2006). Here, a skeletonized map was generated by thinning the mean FA map across participants. The mean FA skeleton was thresholded at FA > 0.2 and then projected onto the normalized FA maps. The same transformation was applied for MD, resulting in voxel-wise FA and MD skeletons for each participant. Using these maps, representing the core of major white matter pathways, we computed mean global FA and MD for each individual.

### 2.5. Brain age prediction

In line with a recent implementation (Kaufmann et al., 2019), age prediction models were trained using XGBoost (extreme gradient boosting) in R (Chen and Guestrin 2016; Chen et al., 2017) based on cortical volume, thickness, and area (Glasser et al., 2016), as well as subcortical volumes (Fischl et al., 2002) as features, totalling 1118 features for each individual (Kaufmann et al., 2019). We trained the models separately for each sex, ensuring that possible sex-related differences in brain aging did not influence the results. Parameters were tuned in a nested cross-validation that estimated the optimal number of model training iterations (settings: nround = 1500, early stopping rounds = 20). The learning rate was set to eta = 0.01 while all other parameters were left as default. To comply with the notion of heterogeneous brain aging we trained two different brain age models, one for the full brain, including all features, and one for cerebellar/ subcortical features alone (Kaufmann et al., 2019). The models were validated with 10-fold cross validation. Finally, the two models were applied to calculate brain age for each individual in the unseen test sample (n=122).

To account for a well-known bias in age prediction we used a described procedure (de Lange & Cole, 2020). First, we calculated the difference between estimated brain age and chronological age. Next, we estimated the association between BAG and age using linear models including age and sex in the models. The beta representing this bias was then subtracted from the estimated age, yielding a corrected brain age for each individual. Chronological age was subtracted from this corrected brain age, and the resulting BAG was used to test for associations with measures of physical activity and capability.

### 2.6. Statistical analysis

Statistical analyses were performed using R, version 3.6.2 (R Core Team, 2019). Data from the StepWatches were analysed with an in-house script, calculating mean (SD) daily activity. Descriptive data are presented as mean (SD) or median (IQR), as appropriate. Due to the use of multidimensional measures of physical activity and capability, and to examine to which degree the measures reflect differentiable components, bivariate associations between measures were examined using Kendall’s tau. The same test of correlation was also used to test for various associations between physical activity, physical capability, sample characteristics, age, and body mass index (BMI). Sex differences in the physical measures were examined using Wilcoxon Rank Sum Test or two-sample t test, as appropriate. To test for interactions between age and sex on the physical measures, Bayes Factor was computed as implemented in the BayesFactor package in R (Richard & Rouder, 2018) and used to compare models with and without the interaction term. To validate the estimation of brain age in the training set, a 10-fold cross validation was used for men and women separately. Further, correlation between estimated and chronological age for the test-set was assessed divided by sex, and finally the association between the BAGS, global FA and MD and age were assessed.

Multiple linear regression analyses were performed using lm in R to test for associations between physical activity and physical capability and BAGs and white matter integrity. For each MRI variable (global and subcortical BAG, global MD and global FA) each of the physical activity and capability variables (daily number steps, grip strength, 10 MWT, and postural control) were included as dependent variables in different linear models, with the MRI measure and age and sex as independent variables. Regression diagnosis for influential cases on the models were performed using Cook’s distance. The results indicated that no cases were overly influential on the results. To explore the association between physical activity and capability and structural brain health further, we ran the regression analysis adding an interaction term between each MRI variable and sex, testing for interaction effect between sex and structural brain health on the physical measures.

Using Fisher z-transformation of the standardized coefficient from the different models, we tested if BAG and DTI measures showed differential associations with the physical health indicators (daily steps, walking speed, hand-grip strength, and postural control). For transparency, we report both uncorrected and corrected p-values. The significance threshold was set at *p* < .05, and for corrected p-values, we employed a Bonferroni correction method. Bayes Factor was also reported alongside p-values for all models in order to quantify the evidence for the null hypothesis, in line with recent recommendations (Keysers et al., 2020).

## 3. Results

### 3.1. Sample characteristics

Table 1 summarizes demographic and clinical characteristics. Average years of education was 15.7 (SD=3.45), ranging from 8 to 27 years. Mean BMI was 24.9 (SD=3.38), ranging from 17.2 to 38.1. Median MMSE score was 29 (IQR= 2), ranging from 24 to 30.

**Table 1.**
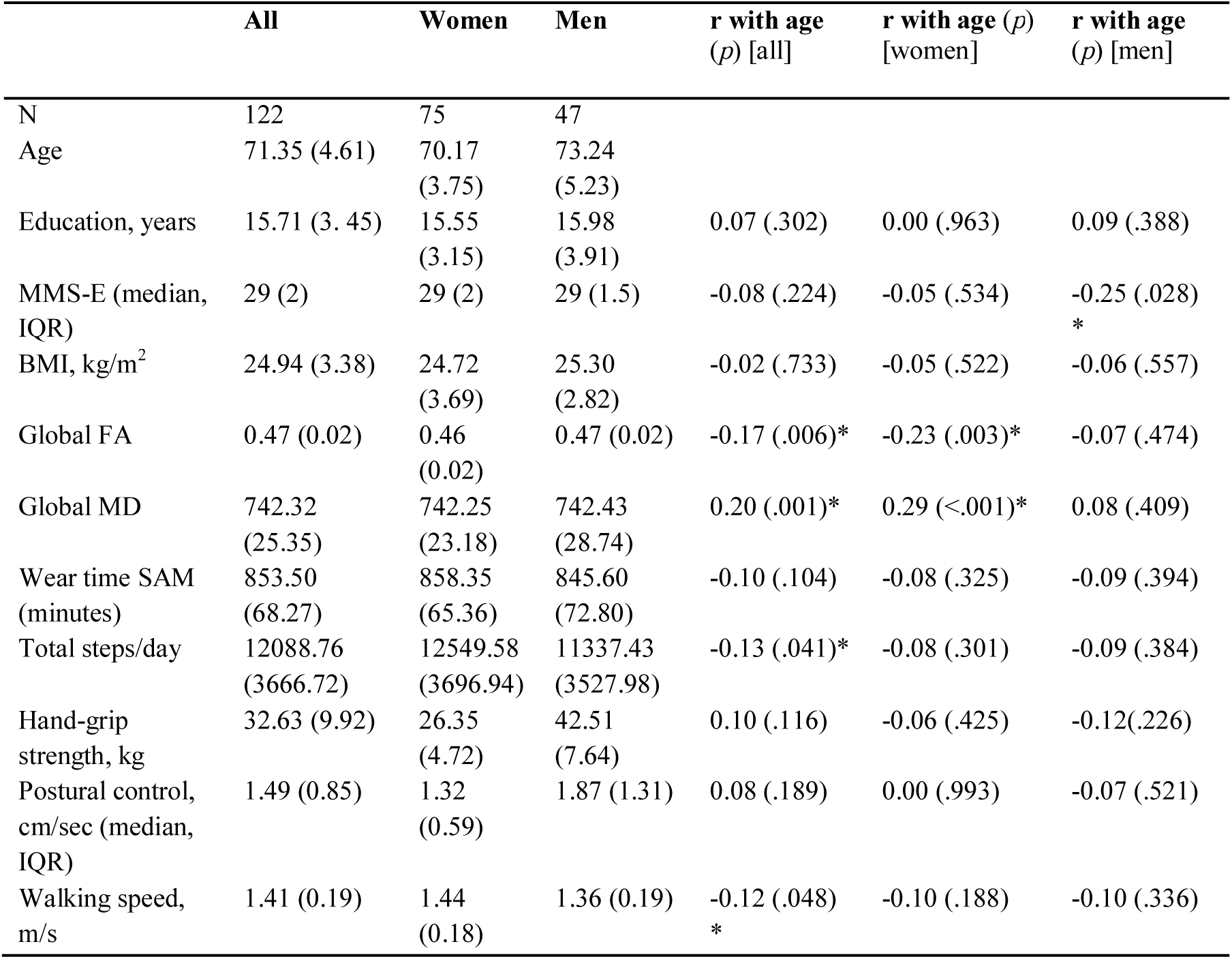
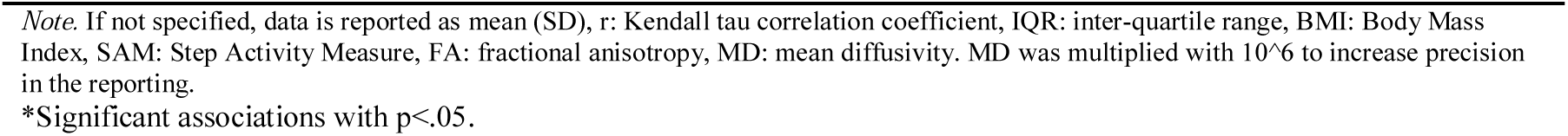
Characteristics of the StrokeMRI sample.

|  | All | Women | Men | r with age<br>(p) [all] | r with age (p)<br>[women] | r with age<br>(p) [men] |
| --- | --- | --- | --- | --- | --- | --- |
| N | 122 | 75 | 47 |  |  |  |
| Age | 71.35 (4.61) | 70.17<br>(3.75) | 73.24<br>(5.23) |  |  |  |
| Education, years | 15.71 (3.45) | 15.55<br>(3.15) | 15.98<br>(3.91) | 0.07 (.302) | 0.00 (.963) | 0.09 (.388) |
| MMS-E (median,<br>IQR) | 29 (2) | 29 (2) | 29 (1.5) | -0.08 (.224) | -0.05 (.534) | -0.25 (.028)<br>* |
| BMI, kg/m<sup>2</sup> | 24.94 (3.38) | 24.72<br>(3.69) | 25.30<br>(2.82) | -0.02 (.733) | -0.05 (.522) | -0.06 (.557) |
| Global FA | 0.47 (0.02) | 0.46<br>(0.02) | 0.47 (0.02) | -0.17 (.006)* | -0.23 (.003)* | -0.07 (.474) |
| Global MD | 742.32<br>(25.35) | 742.25<br>(23.18) | 742.43<br>(28.74) | 0.20 (.001)* | 0.29 (<.001)* | 0.08 (.409) |
| Wear time SAM<br>(minutes) | 853.50<br>(68.27) | 858.35<br>(65.36) | 845.60<br>(72.80) | -0.10 (.104) | -0.08 (.325) | -0.09 (.394) |
| Total steps/day | 12088.76<br>(3666.72) | 12549.58<br>(3696.94) | 11337.43<br>(3527.98) | -0.13 (.041)* | -0.08 (.301) | -0.09 (.384) |
| Hand-grip<br>strength, kg | 32.63 (9.92) | 26.35<br>(4.72) | 42.51<br>(7.64) | 0.10 (.116) | -0.06 (.425) | -0.12(.226) |
| Postural control,<br>cm/sec (median,<br>IQR) | 1.49 (0.85) | 1.32<br>(0.59) | 1.87 (1.31) | 0.08 (.189) | 0.00 (.993) | -0.07 (.521) |
| Walking speed,<br>m/s | 1.41 (0.19) | 1.44<br>(0.18) | 1.36 (0.19) | -0.12 (.048)<br>* | -0.10 (.188) | -0.10 (.336) |

Note. If not specified, data is reported as mean (SD), r: Kendall tau correlation coefficient, IQR: inter-quartile range, BMI: Body Mass Index, SAM: Step Activity Measure, FA: fractional anisotropy, MD: mean diffusivity. MD was multiplied with $10^6$ to increase precision in the reporting.
\*Significant associations with $p < .05$ .

### 3.2. Physical capability and physical activity level

Table 1 summarizes the physical capability and activity measures. Mean hand-grip strength was 32.6 kg (SD=9.92). Men had a significantly greater hand-grip strength (mean 42.5 (SD = 7.64) kg) compared to women (mean 26.3 (SD = 4.7) kg; t = −13.0, *p* = < .001). Median postural control was 1.32 (IQR = 0.59) cm/sec. Women (median 1.41 (IQR = 0.53) cm/s) exhibited significant less postural control than men (median 1.87 (IQR = 1.31) cm/s; W = 807; *p* = < .001, r = −.455). Average walking speed was 1.41 (SD = 0.19) m/s, with women (1.44 (SD = 0.18) m/s) having a significantly higher average speed than men (1.35 (0.19) m/s.; t = 2.48, *p* = .015). The participants had an average of 12088.8 (SD = 3666.72) steps per day, with no significant sex differences (t = 1.8, *p* = 0.08).

Bivariate correlation analysis revealed weak relationships between the various physical test performances, age, and BMI (Fig. 1). In addition, Bayes Factor suggested no evidence of interactions between age and sex on walking speed (BF=0.21, ± 4.34%), daily steps (BF = 0.40, ± 1.9%), hand-grip strength (BF = 0.64, ± 2.13%), or postural control (BF = 0.11, ± 6.82%).

**Fig. 1.**
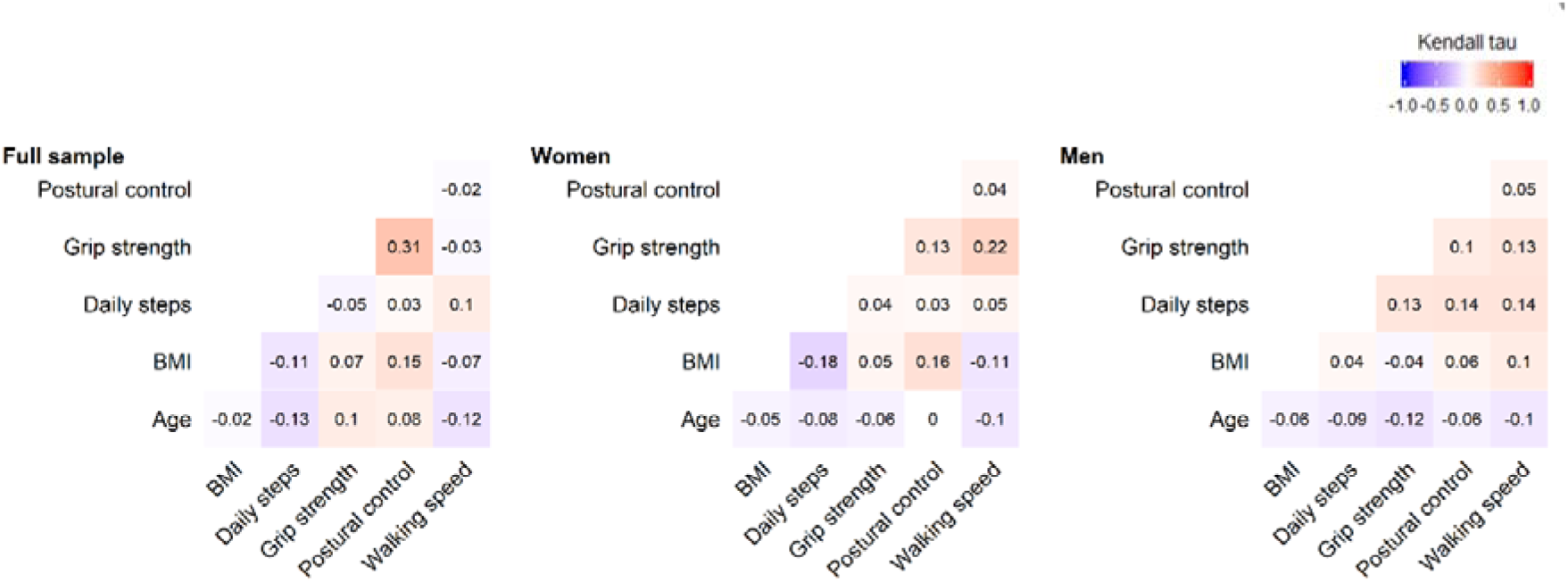
Correlations (Kendall tau) between the physical measurements. Heatmaps showing the associations between the measures of physical capability and activity for the full sample (N = 122), among women (N = 75) and among men (N= 47).

### 3.3. Brain age prediction

In the training set, 10-fold cross validation revealed a correlation of r = 0.90 for both men and women (*p* < .001) between estimated brain age from the full brain model and chronological age (men: MAE = 6.82, RMSE = 8.49, women: MAE = 6.64, RMSE = 8.47). The subcortical model yielded a correlation of r = 0.87 for both men and women (*p* < .001, Men: MAE = 7.49, RMSE = 9.62, Women: MAE = 7.45, RMSE = 9.54). In the test set, the correlation between estimated and chronological age for the full model for men was r = 0.32 (*p* = .002, MAE = 6.44, RMSE = 7.59) and r = 0.34 (*p* < .001, MAE = 6.93, RMSE = 8.53) for women. The subcortical model yielded a correlation of r = 0.28 (*p*= .005, MAE = 7.90, RMSE = 9.39) for men, and r = 0.26 (*p* < .001, MAE = 6.94, RMSE = 8.54) for women.

Fig. 2 shows the association between the BAGs and DTI measures.

**Fig. 2.**
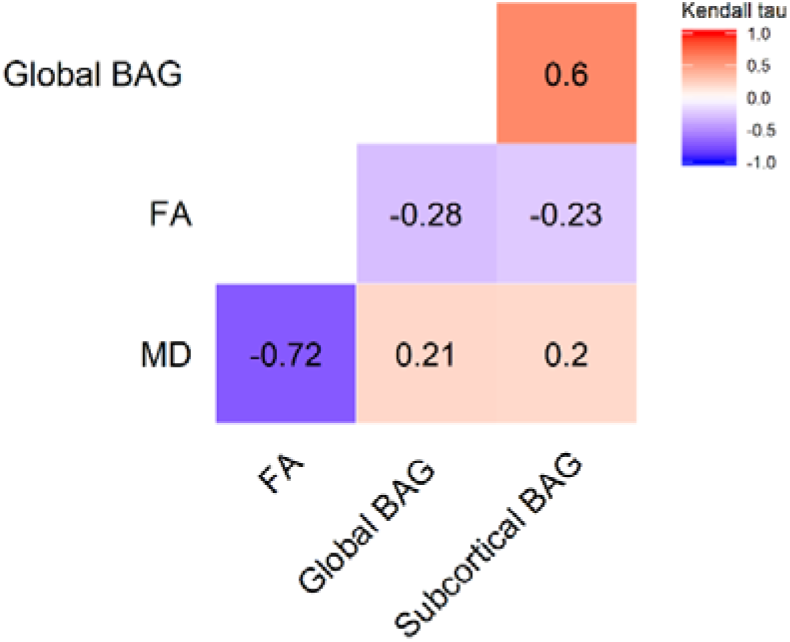
Correlation matrix of the brain imaging measures (Kendall tau) indicating their shared variance.

### 3.4. Association between physical activity, physical capability and BAG

Table 2 summarizes the results from the linear models testing for associations between BAG and measures of physical activity and capability. No significant associations were found after correcting for multiple comparisons, with Bayes Factors generally suggesting equivocal to low evidence for the models not including BAGs compared to the models including BAGs.

**Table 2.** Results of linear models examining the association between physical activity and capability and brain age.

|  | Daily steps |  |  | Walking speed |  |  | Hand-grip strength |  |  | Postural control |  |  |
| --- | --- | --- | --- | --- | --- | --- | --- | --- | --- | --- | --- | --- |
| | $\beta$ | $p$ | BF (error %) | $\beta$ | $p$ | BF (error %) | $\beta$ | $p$ | BF (error %) | $\beta$ | $p$ | BF (error %) |
| Age | -0.19 | **.041\*** | - | -0.15 | .103 | - | -0.13 | .021 | - | -0.08 | .360 | - |
| Sex [Male] | -0.11 | .254 | - | -0.19 | **.039\*** | - | 0.82 | **<.001\*\*** | - | 0.44 | **<.001\*\*** | - |
| Global BAG | -0.07 | .468 | 0.33 ±1.84 | -0.17 | .053 | 1.42 ±2.7 | -0.12 | **.033\*** | 2.20 ±2.95 | -0.13 | .120 | 0.85±1.82 |
| R<sup>2</sup> | 0.08 |  |  | 0.10 |  |  | 0.66 |  |  | 0.23 |  |  |
| F | 2.68 (p=.05) |  |  | 4.34 (p=.006)** |  |  | 76.90 (p<.001)** |  |  | 10.38 (p<.001)** |  |  |
| Age | -0.2 | **.037\*** | - | -0.15 | .101 | - | -0.13 | **.024\*** | - | -0.08 | .361 | - |
| Sex [Male] | -0.12 | .195 | - | -0.2 | **.030\*** | - | 0.83 | **<.001\*\*** | - | 0.44 | **<.001\*\*** | - |
| Subcortical BAG | -0.13 | .141 | 0.34 ±1.83 | -0.19 | **.033\*** | 2.05 ±2.06 | -0.08 | .156 | 0.72 ±2.62 | -0.12 | .161 | 0.71 ±1.41% |
| R<sup>2</sup> | 0.15 |  |  | 0.11 |  |  | 0.66 |  |  | 0.21 |  |  |
| F | 3.27 (p=.024)* |  |  | 4.64 (p=.004)** |  |  | 74.41 (p<.001)** |  |  | 10.19 (p<.001)** |  |  |

*Note:* $\beta$ = standardized coefficients. Bayes Factor (BF) was estimated using BayesFactor package in R, and represent the evidence of the full model against null model. Null model: dependent variable ~ age + sex.
\*Significant associations with p<.05
\*\*Significant associations after Bonferroni correction.

Models including subcortical BAG by sex interactions revealed a significant main effect of daily steps (β = −0.35, *p* = .003), indicating lower subcortical BAG in people with a higher number of daily steps, and a significant interaction between subcortical BAG and sex (β = 0.34, *p* = .003), with Bayes Factor suggesting strong evidence for the model including the interaction term (BF = 16.51, ± 1.63%) (Supplementary Table 4). Follow-up analyses revealed a significant association between subcortical BAG and number of steps among women (β = −0.34, *p* = .003), but not among men (β = 0.20, *p* = 0.172) (Fig. 3). For the remaining models, including the interaction term did not change the results, with Bayes Factor in general suggesting equivocal to low evidence for the models not including interaction effects.

**Fig. 3.**
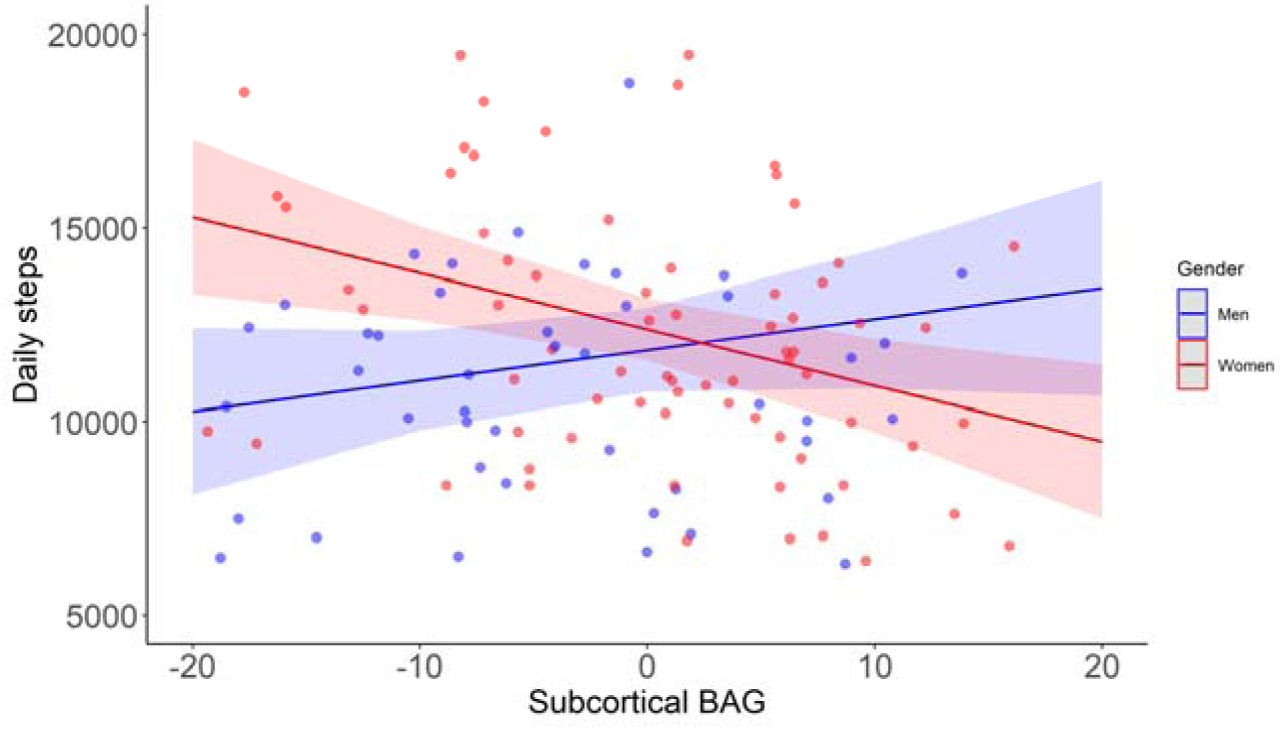
Associations between daily steps and subcortical BAG among men and women.

### 3.5. Association between physical activity, physical capability and DTI measures

Supplementary Fig. 2 shows the association between age and DTI measures. Global FA (r_τ_ = −0.17, *p* < .006, BF = 7.85, ± 0%) and MD (r_τ_ = 0.20, *p* < .001, BF = 29.45, ± 0%) were significantly associate with age.

Table 3 summarizes the results from the linear models testing for associations between the DTI measures and the various measures of physical activity and capability. After correction for multiple comparisons, linear models revealed a significant positive association between global FA and walking speed (β = 0.25, *p* = .006, BF = 7.5, ± 2.72%), with the model including FA being preferred by Bayes Factor. The results are indicating higher average pace with higher FA. For walking speed, Bayes Factor also suggested moderate evidence for the model including global MD compared to the model including only age and sex, indicating higher walking speed with lower MD. This association did not remain significant after correction for multiple comparisons (β = −0.24, *p* = .01, BF = 5.05 ± 2.73%). No other significant associations were found, with Bayes Factors generally suggesting equivocal to low evidence for the models not including DTI-measures compared to the models including DTI-measures, and with similar results when adding the interactions with sex (Supplementary Table 5).

**Table 3.**
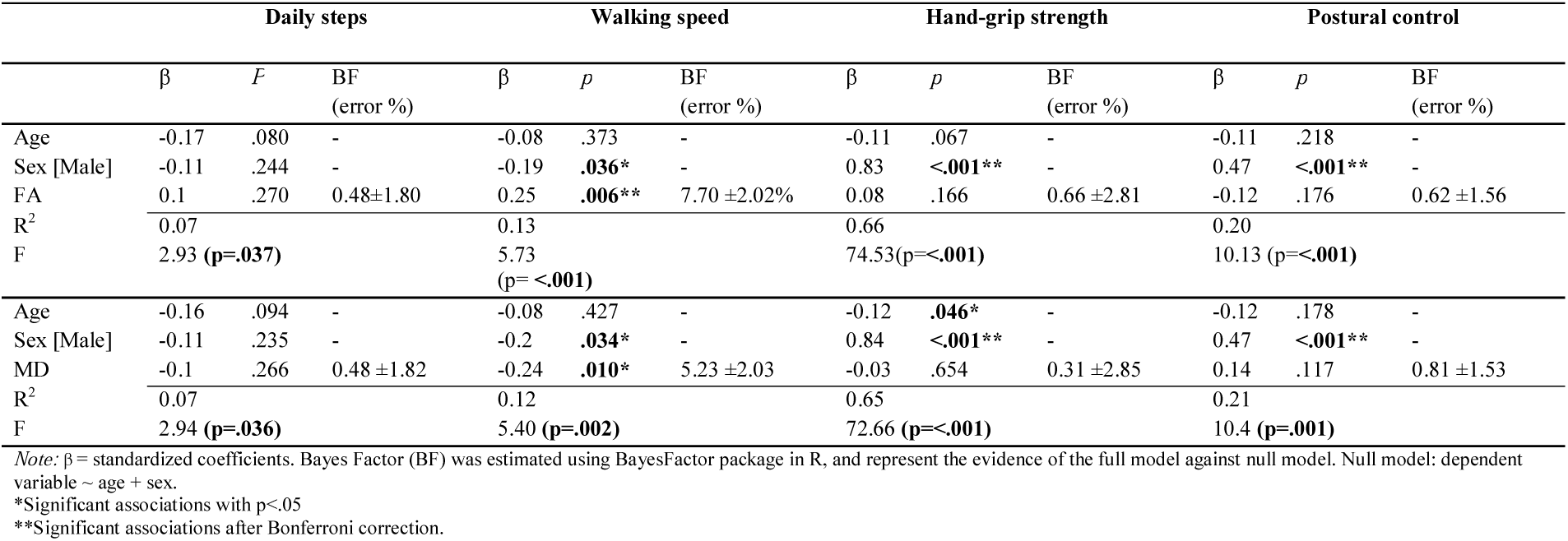
Results of linear models examining the association between physical activity and capability and DTI measures.

### 3.6. Differences in sensitivity between the various imaging results

Fisher z-transformation indicated differences in the associations between walking speed and FA compared to global BAG (t = 3.35, *p* = .001), subcortical BAG (t = 3.48, *p* = .001), and MD (t = 3.8, *p* < .001). No other robust differences were found in the associations between physical activity or physical capability between the different imaging modalities (Supplementary Table 6).

## 4. Discussion

The purpose of the current study was to examine the association between objective measures of physical activity level and capability, and two complementary structural MRI based indicators of brain health in healthy older adults. The study focuses on identifying lifestyle-factors with potential dissimilar protecting effect against gray and white matter changes in the aging brain. Our study indicates a positive association between daily steps and a younger appearing brain based on the subcortical model in women, but not in men, with Bayesian analyses strongly supporting an association. Moreover, our results indicate that a higher walking speed is associated with more coherent white matter structure as indicated by global FA, with Bayesian analysis suggesting moderate evidence for the association.

### 4.1. Daily activity level and brain health

Based on the assumption that physical activity promotes brain health in older adults, we hypothesized that people with higher levels of physical activity would show less evident brain aging than their peers who are less physically active. In line with this, our analyses revealed higher subcortical BAG in participants with fewer daily steps. These results support and extend previous studies (Erickson et al., 2014; Hamer et al., 2018) by demonstrating that the relationship persists when using brain age prediction and a sensor-based measure of activity level. The latter is also considered to provide a more valid proxy of activity level than questionnaires. One explanation for this is that older adults get more daily physical activity from low-intensity activity (Dyrstad et al., 2014), which may be more difficult to recall than hours at the gym or other specific events (Guo et al., 2019).

Our findings of younger appearing brains in people with higher physical activity levels could reflect various biological mechanisms. Physical activity has been reported to influence the levels of neurotropic factors (brain-derived neurotrophic factor (BDNF), Insulin-like Growth Factor 1 (IGF-1), and Vascular Endothelial Growth Factor (VEGF) (Lee et al., 2014), which are central for preventing deterioration of neurons. Physical activity also has beneficial effects for the neurovascular system, as measured using cerebral blood flow and perfusion (Klenk et al., 2013). Supporting the link between cardiovascular and brain health, higher brain age in middle-aged and elderly people with cardiovascular risk factors such as high blood pressure, alcohol intake, and stroke risk score were recently reported, with blood pressure showing a stronger association with white matter compared to gray matter (de Lange et al., 2020a).

In contrast to previously reported associations between white matter measures and sensor-measured physical activity (Burzynska et al., 2014; Tian et al., 2015), our analysis revealed no significant associations between physical activity and global BAG or white matter DTI. Sample characteristics such as the average and variance in physical activity may partly explain the discrepancies in results between studies. Our participants had an average of 12,088 steps per day, which is high compared to normative data for adults (mean 2000-9000 steps per day) (Tudor-Locke et al., 2009). In addition, while previous studies assessed regional effects, our results are based on more global measures of BAG and white matter DTI. Finally, we used total daily steps as a proxy of overall physical activity, and did not include intensity or temporal aspects of daily physical activity, which are likely relevant factors.

The association between subcortical BAG and daily activity level was only present in women. This is in line with a former study suggesting an association between total daily walking with larger hippocampal volume, but only in women (Varma et al., 2015). The results might be related to the postmenopausal hormonal changes (Dalal & Agarwal, 2015). A recent study demonstrated that higher estimated levels of sex-hormone exposure was associated with higher brain age in women (de Lange et al., 2020b). In addition, an interaction between hormone replacement therapy and fitness on age related decline in gray matter volume has been demonstrated (Erickson et al., 2007), indicating increased neuroprotective effect of fitness in combination with hormone replacement treatment for post-menopausal women. Our current results might also be related to the population studied, but there was no sex difference in activity level or any other obvious sex differences present. The results, though in line with previous publications (Barha et al., 2017; Liu-Ambrose et al., 2018; Varma et al., 2016), must be interpreted carefully and warrant replication in future studies.

### 4.2. Physical capability and brain health

In the present study we hypothesized that indicators of physical capability such as grip strength, walking speed, and postural control would associate with structural MRI based indicators of brain health in healthy older adults. In contrast to a previous study on participants aged 73 years (Cole & Franke, 2017), we found no significant associations between walking speed and global or subcortical BAG. However, supporting the hypothesis, the results demonstrated a positive association between white matter FA and walking speed. These results are in line with a previously reported association between white matter measures and a latent variable for physical fitness created from grip strength, forced respiratory volume, and 6-metre walk time, in participants aged 76 years (Ritchie et al., 2017). The association between coherence of white matter and walking speed could be of importance because of the possibility that structural brain changes may represent a mediator between physical capability and cognitive function in senescence. By facilitating swift and synchronised information flow, brain white matter microstructure has been demonstrated to provide critical support for cognitive functions (Strömmer et al., 2020). Moreover, maintaining white matter integrity has formerly been identified as an important predictor for successful cognitive aging (Kennedy & Raz, 2009). The current study design does not allow for causal inference, and our results do not necessarily imply that improving walking speed in older age will contribute to a better brain health, or vice versa. Supporting a possible causal association, a recent large-scale study utilizing data from UK Biobank identified 70 independent genetic loci with significant associations with self-reported walking speed. Approximately 10% of the variance in self-reported walking speed was attributed to individual differences in common genetic variants, and significant genetic correlations were reported between self-reported walking speed and cardiometabolic, respiratory and psychiatric traits, educational attainment and mortality.

Further, follow-up Mendelian randomization analyses, which allows for causal inference, suggested that increasing walking pace decreases cardiometabolic risk, in line with current public health advice (Timmins et al., 2020). Moreover, a number of studies have reported that walking speed is a strong predictor of mortality (Cooper et al., 2010; Ganna & Ingelsson, 2015) and a relevant index of “vital aging” (Vermeulen et al., 2011). The results of the present study substantiate the possible importance of walking speed as a target in public health interventions, and as a possible index of white matter brain health in older adults.

In addition to walking speed, hand-grip strength and postural control has been suggested as markers of healthy aging due to multiple associations with measures of health (Bohannon, 2019; Vermeulen et al., 2011). Both strength and postural control dependent on mechanical contributions from both the muscles-, skeletal, and joint systems (Granacher et al., 2008), but also visual, vestibular, haptic and proprioceptive information are critical in specific for maintaining postural control (Alcock et al., 2018). Integration of this information has been suggested to be partly related to the structure and function of the brain, and potentially be affected by the aging process in the brain with following reduction of function (Sullivan et al., 2009). In contrast to this and other previous work (Cole et al., 2018), our analyses revealed no significant associations between brain MRI and hand-grip strength in a presumably healthy sample. Former publications including mainly clinical populations have characterized postural control as a “whole brain phenomenon”, but also highlighted cerebellum as an important region (Surgent et al., 2019). Our results indicated no significant associations between subcortical or global BAG or DTI measures of brain integrity and postural control. For FA and MD, this was in line with a previous study reporting weak associations in the postural control association with regional FA, and a lack of association with MD (Massa et al., 2019). We are not aware of previous studies testing for associations between subcortical BAG, including the cerebellum, or global BAG and postural control measured with a forced pressure platform. However, in relation to former studies highlighting cerebellum as important structure for postural control, this is also suggested to be a region more implicated in clinical populations than in healthy ones (Surgent et al., 2019), and this might substantiate the findings in the present study. The neurobiological underpinning in the brain of variation in postural control in healthy older adults remains uncertain.

### 4.3. Methodological considerations

The strengths of our study include objectively measured physical activity, several objective and sensitive tests of physical capability, and the advanced multimodal imaging approach.

Some limitations should also be emphasized. The study sample was relatively homogenous and high functioning in terms of level of education and physical activity, which may have limited the sensitivity. With increasing age, it is conceivable that a more selective and less representative part of the population volunteers for a study including an extensive protocol including both multimodal MRI and physical and neuropsychological tests. Further studies are needed to test the generalisability to other populations.

The cross-sectional design does not permit inference about brain changes. Longitudinal studies covering a larger part of the lifespan are required to explore the dynamics of the associations, for example to which degree the effects of physical activity vary across the lifespan, and to which degree early intervention may protect against age-related decline years or decades later.

### 4.4. Conclusions

In conclusion, we have demonstrated that different markers of brain white matter structure and brain aging are associated with objectively measured daily activity and physical capability. The strongest associations were found between subcortical BAG and daily physical activity, but only for women, and between global white matter FA and walking speed. While larger longitudinal studies are needed to explore potential causal and long-term effects of physical activity on brain aging, our results suggest that combining multimodal measures of brain structure provides complementary information and show dissociable associations with physical activity and capability in elderly healthy adults.

## Data Availability

The StrokeMRI data incorporated in this paper is not available online.
The rest of the data incorporated in this work were gathered from the following open access datasets:
1. The Cambridge Centre for Ageing and Neuroscience (Cam-CAN)
2. The Dallas Lifespan Brain Study (DLBS)
3. OpenfMRI database
4. IXI
5. OASIS
6. Southwest University Adult Lifespan Dataset (SALD)

https://camcan-archive.mrc-cbu.cam.ac.uk/dataaccess/

http://fcon_1000.projects.nitrc.org/indi/retro/dlbs.html

https://openfmri.org/

https://openneuro.org/datasets/ds000222/versions/00001

http://brain-development.org/ixi-dataset/

http://www.oasis-brains.org/

http://fcon_1000.projects.nitrc.org/

http://fcon_1000.projects.nitrc.org/indi/retro/sald.html

## Acknowledgments and funding

We are very grateful to all those who have participated in the StrokeMRI study. The study was funded by the Research Council of Norway [249795, 248238, 276082], the South-Eastern Norway Regional Health Authority [2014097, 2015044, 2015073, 2018037, 2019107, 2020086], the Norwegian ExtraFoundation for Health and Rehabilitation [2015/ FO5146], the European Research Council under the European Union’s Horizon 2020 research and Innovation program [ERC StG Grant 802998], Sunnaas Rehabilitation Hospital HT, and the Department of Psychology, University of Oslo. Funding of the data sources used in the training set is summarized in Supplementary material Table 2.

## Author contribution

**Anne-Marthe Sanders:** Conceptualization, Software, Formal analysis, Investigation, Data Curation, Writing - Original Draft, Writing - Review & Editing, Visualization, **Geneviève Richard:** Conceptualization, Investigation, Data Curation, Writing - Review & Editing, **Knut Kolskår:** Conceptualization, Software, Investigation, Data Curation, Writing - Review & Editing, **Kristine M. Ulrichsen:** Investigation, Data Curation, Writing - Review & Editing, **Tobias Kaufmann:** Software, Validation, Formal analysis, Writing - Review & Editing, **Dag Alnæs:** Conceptualization, Writing - Review & Editing, **Dani Beck:** Writing - Review & Editing, **Erlend S. Dørum:** Conceptualization, Investigation, Writing - Review & Editing, **Ann-Marie G. de Lange:** Writing - Review & Editing, **Jan Egil Nordvik:** Conceptualization, Writing - Review & Editing, Supervision, Funding acquisition, **Lars T. Westlye:** Conceptualization, Software, Formal analysis, Resources, Writing - Original Draft, Writing - Review & Editing, Supervision, Project administration, Funding acquisition

## Web references

http://surfer.nmr.mgh.harvard.edu (accessed 21 January 2021)

https://fsl.fmrib.ox.ac.uk/fsl (accessed 21 January 2021)

https://fsl.fmrib.ox.ac.uk/fsl/fslwiki/eddy (accessed 21 January 2021)

https://fsl.fmrib.ox.ac.uk/fsl/fslwiki/topup (accessed 21 January 2021)

