## Supplementary material for "Linking objective measures of physical activity and capability with brain structure in healthy community dwelling older adults"

**Authors:** Anne-Marthe Sanders^a,b,c,*^, Geneviève Richard^a,^ , Knut Kolskår^a,b,c^, Kristine M. Ulrichsen^a,b,c^, Tobias Kaufmann^a,d^, Dag Alnæs^a,e^, Dani Beck^a,b,c^, Erlend S. Dørum^a,b,c^, Ann-Marie G. de Lange^a,b,f^, Jan Egil Nordvik^g^ , Lars T. Westlye^a,b,h^

*^a^ NORMENT, Division of Mental Health and Addiction, Oslo University Hospital & Institute of Clinical Medicine, University of Oslo, Norway*

*^b^ Department of Psychology, University of Oslo, Norway*

*^c^ Sunnaas Rehabilitation Hospital HT, Nesodden, Norway*

*^d^ Department of Psychiatry and Psychotherapy, University of Tübingen, Germany*

*^e^ Bjørknes college, Oslo, Norway*

*^f^ Department of Psychiatry, University of Oxford, Warneford Hospital, Oxford, UK*

*^g^ CatoSenteret Rehabilitation Center, Son, Norway*

*^h^ KG Jebsen Center for Neurodevelopmental Disorders, University of Oslo, Norway*

| Supplementary Table 1. Descriptions of the training cohort. \| \| \|; **Training cohort** \| **N** \| **Age (years):** mean ± SD \| **Sex (women/ men)**; The Cambridge Centre for Ageing and Neuroscience (Cam-CAN) \| 648 \| 54.2±18.6 \| 329/319; Dallas Lifespan Brain Study (DLBS) \| 311 \| 54.3±20.0 \| 194/117; OpenfMRI/ ds000222 \| 79 \| 44.4±20.1 \| 41/38; IXI \| 562 \| 48.6±16.5 \| 312/250; Open Access Series of Imaging Studies (OASIS) \| 314 \| 45.0±23.9 \| 196/118; Southwest University Adult Lifespan Dataset (SALD) \| 493 \| 45.2±17.4 \| 307/186 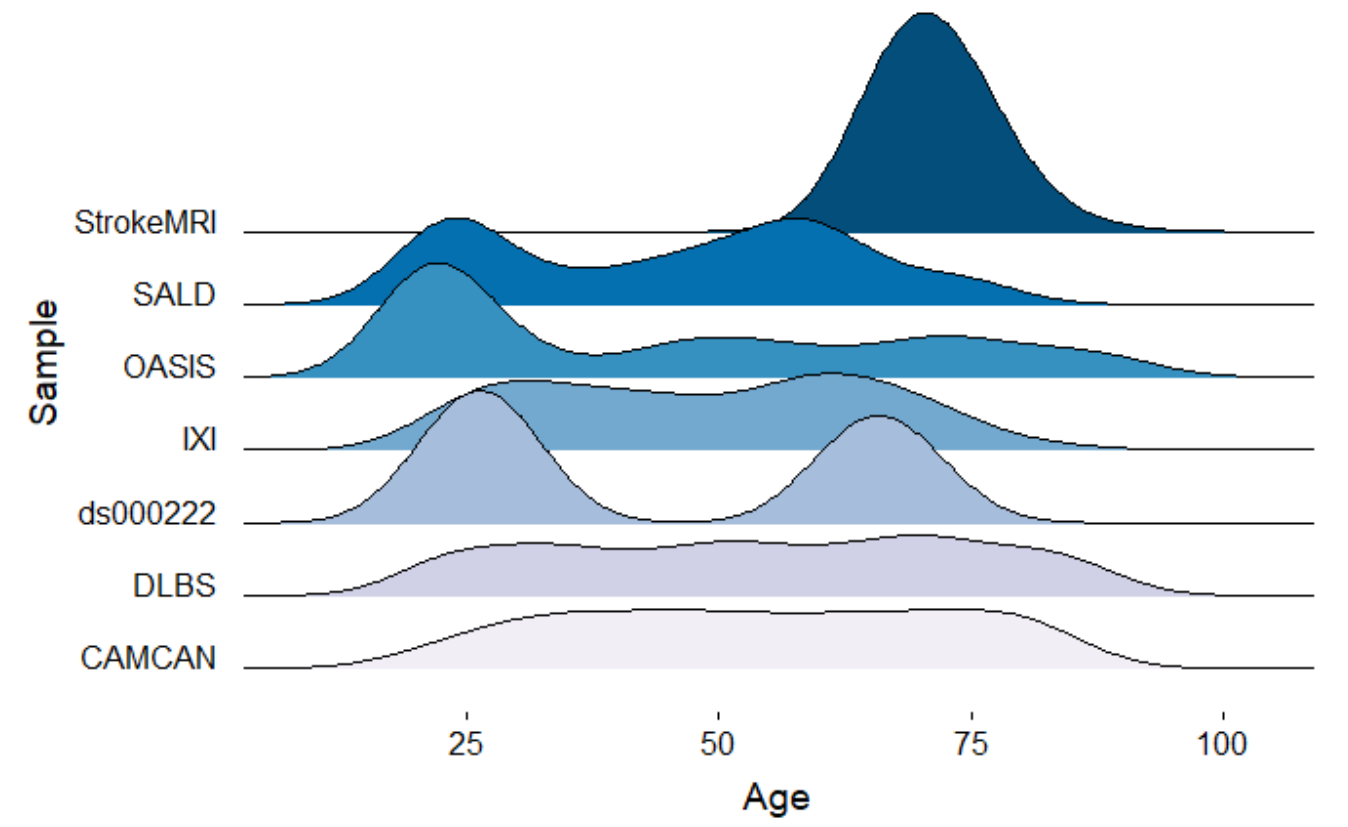 **Supplementary Fig. 1.** Age distribution for all samples. **Supplementary Table 2.** Summary of the training-cohorts. | | | |
| --- | --- | --- | --- |
| **Cohort** | **Source** | **Information** | **Reference** |
| Cam-CAN | https://camcan-archive.mrc-cbu.cam.ac.uk/dataaccess/ | The Cambridge Centre for Ageing and Neuroscience (Cam-CAN) provided the data. Cam-CAN was funded from the Biotechnology and Biological Sciences Research Council ([BBSRC](http://www.bbsrc.ac.uk/), grant number BB/H008217/1), the Medical Research Council (MRC) [Cognition & Brain Sciences Unit (CBU)](http://www.mrc-cbu.cam.ac.uk/), the European Union Horizon 2020 [LifeBrain project](https://www.lifebrain.uio.no/) and University of Cambridge, UK. | (Shafto et al., 2014; Taylor et al., 2017) |
| DLBS | http://fcon_1000.projects.nitrc.org/indi/retro/dlbs.html | The data-collection for the Dallas Lifespan Brain Study (DLBS) was supported by the National Institutes of Health grants R37 AG006265 to D.C.P., R21 AG034318 to H.L., R01 AG033106 to H.L., and R01 MH084021 to H.L. | (Lu et al., 2011) |
| OpenfMRI/ ds000222 | https://openfmri.org/  https://openneuro.org/datasets/ds000222/versions/00001 | The data was obtained from the OpenfMRI database. Its accession number is ds000222 | (FitzGerald et al., 2017; Gorgolewski et al., 2017; Poldrack et al., 2016) |
| IXI | http://brain-development.org/ixi-dataset/ |  | (Liu et al., 2017) |
| OASIS | http://www.oasis-brains.org/ | The study was supported by grants P50 AG05681, P01 AG03991, R01 AG021910, P50 MH071616, U24 RR021382, R01 MH56584 | (Buckner et al., 2004; Fotenos et al., 2005) |
| SALD | http://fcon_1000.projects.nitrc.org/  http://fcon_1000.projects.nitrc.org/indi/retro/sald.html | Southwest University Adult Lifespan Dataset (SALD) | (Wei et al., 2018) |
| *Note.* In the training-cohorts the collection of data was not completed, as follows, the number of subjects in the reference publication does not match the current study. | | | |

**Supplementary Table 3.** Image acquisition for the training-samples.

| Cohort/ scanner, head coil | T1 parameters | Reference |
| --- | --- | --- |
| Cam-CAN/ 3T MRC-CBSU, 32-channel head coil | Sequence: Magnetization prepared rapid acquisition of gradient echo (MPRAGE),  Acquisition-time: 4.32 minutes, TE/TR= 2.99 ms/ 2250 ms, FOV= 256mm x 240mm x 192mm, FA: 9^ο^, VS= 1x1x1 mm^3^ | (Shafto et al., 2014) |
| DLBS/ 3T Philips Medical System, 8-channel head coil | Sequence: MPRAGE, Acquisition-time: 3.57 minutes, TE/ TR=3.7 ms/ 8.1 ms, FA: 18^ο^, VS= 1x1x1 mm^3^ | (Lu et al., 2011) |
| OpenfMRI/ ds000222/ Siemens 3T Trio Magnetom | Sequence: MPRAGE  TE/ TR= 2.34 ms/ 1550 ms, FA: 9^ο^, VS= 1x1x1 mm^3^ | (FitzGerald et al., 2017) |
| IXI/ Philips 1.5T, GE 1.5T and a Philips 3T | Sequence: MPRAGE  Parameters for Philips 1.5T: TE/ TR = 4.6 ms/ 9.8 ms, FA: 8°  Parameter for Philips 3T: TE/ TR = 4.6 ms/ 9.6 ms, FA: 8°.  Parameters for GE 1.5T: not available. | (Liu et al., 2016) |
| OASIS/ 1.5T Siemens Vision | Sequence: MPRAGE,  Acquisition-time: 6.6 minutes, TE/ TR= 4 ms/ 9.7 ms, FOV = 256 × 256, FA: 10°, VS: 1x1x1 mm^3^ | (Buckner et al., 2004) |
| SALD/ 3.0T SiemensTrio | Sequence: MPRAGE  TE/ TR=2.52/ 1.9 ms, FA: 90°, VS: 1x1x1 mm^3^ | (Wei et al., 2018) |

**Supplementary Table 4.** Results of regression analysis examining the association between physical activity and physical capability and BAG, including interaction term of sex.

|  | **Daily steps** | | | **Walking speed** | | | **Hand-grip strength** | | | **Postural control** | | |
| --- | --- | --- | --- | --- | --- | --- | --- | --- | --- | --- | --- | --- |
|  | β | *p* | BF  (error %) | β | *p* | BF  (error %) | β | *p* | BF  (error %) | β | *p* | BF  (error %) |
| Age | -0.18 | .060 |  | -0.15 | .122 |  | -0.13 | **.027*** |  | -0.1 | .263 |  |
| Sex [Male] | -0.09 | .328 |  | -0.19 | **.048*** |  | 0.83 | **<.001**** |  | 0.42 | **<.001**** |  |
| Global BAG | -0.14 | .201 |  | -0.2 | .059 |  | -0.14 | **.04*** |  | -0.04 | .701 |  |
| Sex [Male]*Global BAG | 0.13 | .228 | 0.61 ±2.67 | 0.06 | .593 | 0.36±3 | 0.04 | .589 | 0.35 ±1.53 | -0.16 | .107 | 1.01± 1.97 |
| R^2^ | 0.08  2.39  (*p*=.055) | |  | 0.10  3.31 **(*p*=.013)** | |  | 0.66  57.40 **(*p*=<.001)** | |  | 0.23  8.55 **(*p*=<.001)** | |  |
| F |  |  |  |  |  |  |  |  |  |  |  |  |
| Age | -0.19 | **.043*** |  | -0.15 | .105 |  | -0.13 | .025 |  | -0.08 | .347 |  |
| Sex [Male] | -0.07 | .453 |  | -0.19 | **.045*** |  | 0.83 | **<.001**** |  | 0.42 | **<.001**** |  |
| Subcortical BAG | -0.35 | **.002**** |  | -0.24 | **.037*** |  | -0.09 | .204 |  | -0.04 | .736 |  |
| Sex[Male]*Subcortical BAG | 0.35 | **.003**** | 16.51±1.63 | 0.08 | .475 | 0.37±1.92 | 0.02 | .772 | 0.32 ±2.49 | -0.13 | .242 | 0.62±1.95 |
| R2 | 0.15  4.93  **(*p*=.001)** | |  | 0.11  3.59 **(*p*=.008)** | |  | 0.66  55.39  **(*p*=<.001)** | |  | 0.21  8.01  **(*p*=<.001)** | |  |
| F |  |  |  |  |  |  |  |  |  |  |  |  |

*Note:* β = standardized coefficients. Bayes Factor (BF) represent the evidence of the full model against null model. Null model: dependent variable ~ age + sex + imaging data.

*Significant associations with p<.05

**Significant associations after Bonferroni correction.

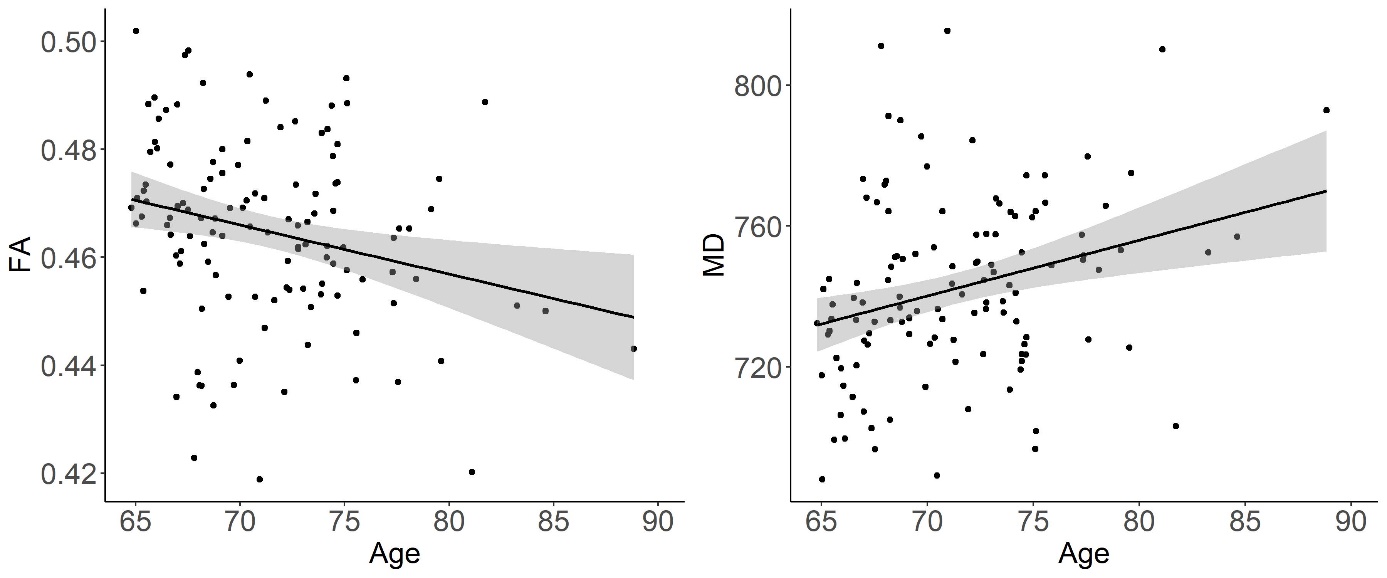

**Supplementary Fig. 2.** Scatterplots of the associations between age and FA and MD. MD was multiplied with 10^6 to increase precision in the reporting.

**Supplementary Table 5.** Results of regression analysis examining the association between physical activity and physical capability and white matter integrity, including interaction term of sex.

|  | **Daily steps** | | | **Walking speed** | | | **Hand-grip strength** | | | **Postural control** | | |
| --- | --- | --- | --- | --- | --- | --- | --- | --- | --- | --- | --- | --- |
|  | β | *p* | BF  (error %) | β | *p* | BF  (error %) | β | *p* | BF  (error %) | β | *p* | BF  (error %) |
| Age | -0.17 | .077 | - | -0.09 | .370 | - | -0.11 | .072 | - | -0.1 | .256 | - |
| Sex [Male] | -0.98 | .696 | - | -0.6 | .797 | - | 1.47 | .32 | - | 4.36 | **.05*** | - |
| FA | 0.07 | .542 | - | 0.23 | .056 | - | 0.1 | .19 | - | 0.02 | .869 | - |
| Sex [Male]* FA | 0.87 | .729 | 0.33 ±2.12% | 0.41 | .862 | 0.30 ±1.75% | -0.64 | .664 | 0.34 ±2.91% | -3.9 | .079 | 1.30±2%* |
| R^2^ | 0.07  2.21 (p=.071) | |  | 0.13  4.27 **(*p*=.003)** | |  | 0.66  55.4 **(*p*=<.001)** | |  | 0.23  8.52 **(*p*=<.001)** | |  |
| F |  |  |  |  |  |  |  |  |  |  |  |  |
| Age | -0.17 | .09 | - | -0.08 | .389 | - | -0.12 | .051 | - | -0.11 | .233 | - |
| Sex [Male] | 0.79 | .768 | - | 1.72 | .491 | - | 0.16 | .92 | - | -3.6 | .127 | - |
| MD | -0.07 | .556 | - | -0.17 | .184 | - | -0.05 | .537 | - | -0.01 | .951 | - |
| Sex[Male]* MD | -0.9 | .736 | 0.34 ±2.12% | -1.92 | .443 | 0.39 ±1.77% | 0.68 | .671 | 0.35±3.03% | 4.07 | .084 | 1.22±1.99 |
| R2 | 0.07  2.12 (*p*=.072) | |  | 0.13  4.18 **(*p*=.003)** | |  | 0.65  54.16 **(*p*=<.001)** | |  | 0.23  8.69 **(*p*=<.001)** | |  |
| F |  |  |  |  |  |  |  |  |  |  |  |  |

*Note:* β = standardized coefficients. Bayes Factor (BF) represent the evidence of the full model against null model. Null model: dependent variable ~ age + sex.

*Significant associations with p<.05

**Significant associations after Bonferroni correction.

**Supplementary Table 6.** Comparison of the standardized coefficient from the linear models

|  | **Test** | **t-value** | ***p*** |
| --- | --- | --- | --- |
| **Daily steps** | Global BAG; Subcortical BAG | -0.54 | 0.593 |
|  | Global BAG; MD | -0.31 | 0.754 |
|  | Global BAG; FA | 1.3 | 0.195 |
|  | Subcortical BAG; MD | 0.21 | 0.837 |
|  | Subcortical BAG; FA | 1.82 | 0.071 |
|  | MD; FA | 1.57 | 0.118 |
| **Hand-grip strength** | Global BAG; Subcortical BAG | 0.5 | 0.617 |
|  | Global BAG; MD | 1.16 | 0.248 |
|  | Global BAG; FA | 2.5 | 0.014 |
|  | Subcortical BAG; MD | 0.67 | 0.506 |
|  | Subcortical BAG; FA | 2 | 0.048 |
|  | MD; FA | 1.3 | 0.197 |
| **Postural control** | Global BAG; Subcortical BAG | 0.1 | 0.918 |
|  | Global BAG; MD | 2.22 | 0.028 |
|  | Global BAG; FA | 0.11 | 0.91 |
|  | Subcortical BAG; MD | 2.11 | 0.037 |
|  | Subcortical BAG; FA | 0.01 | 0.991 |
|  | MD; FA | -2.08 | 0.04 |
| **Walking speed** | Global BAG; Subcortical BAG | -0.14 | 0.885 |
|  | Global BAG; MD | -0.5 | 0.615 |
|  | Global BAG; FA | 3.35 | **0.001**** |
|  | Subcortical BAG; MD | -0.36 | 0.719 |
|  | Subcortical BAG; FA | 3.48 | **0.001**** |
|  | MD; FA | 3.8 | **>0.001**** |

*Significant associations with p<.05

**Significant associations after Bonferroni correction.
